# The extracellular vesicle transcriptome provides tissue-specific functional genomic annotation relevant to disease susceptibility in obesity

**DOI:** 10.1101/2024.11.18.24317277

**Authors:** Emeli Chatterjee, Michael J. Betti, Quanhu Sheng, Phillip Lin, Margo P Emont, Guoping Li, Kaushik Amancherla, Worawan B Limpitikul, Olivia Rosina Whittaker, Kathy Luong, Christopher Azzam, Denise Gee, Matthew Hutter, Karen Flanders, Parul Sahu, Marta Garcia-Contreras, Priyanka Gokulnath, Charles R. Flynn, Jonathan Brown, Danxia Yu, Evan D. Rosen, Kendall Van-Keuren Jensen, Eric R. Gamazon, Ravi Shah, Saumya Das

**Author notes:** These authors contributed equally (Emeli Chatterjee, Michael J. Betti, Quanhu Sheng). These authors jointly supervised the work (Eric R. Gamazon, Ravi Shah, Saumya Das).

## Abstract

We characterized circulating extracellular vesicles (EVs) in obese and lean humans, identifying transcriptional cargo differentially expressed in obesity. Since circulating EVs may have broad origin, we compared this obesity EV transcriptome to expression from human visceral adipose tissue derived EVs from freshly collected and cultured biopsies from the same obese individuals. Using a comprehensive set of adipose-specific epigenomic and chromatin conformation assays, we found that the differentially expressed transcripts from the EVs were those regulated in adipose by BMI-associated SNPs from a large-scale GWAS. Using a phenome-wide association study of the regulatory SNPs for the EV-derived transcripts, we identified a substantial enrichment for inflammatory phenotypes, including type 2 diabetes. Collectively, these findings represent the convergence of the GWAS (genetics), epigenomics (transcript regulation), and EV (liquid biopsy) fields, enabling powerful future genomic studies of complex diseases.

## INTRODUCTION

The search for novel targets in human obesity—a *sine qua non* precursor for contemporary metabolic diseases^1^—has fuelled an array of population-based molecular studies over the last decade^2–8^. In this space, human genetics and accompanying multiomic techniques have identified and reproduced genomic loci^2–3^ and signatures of obesity^9^, suggesting an important role for genetic susceptibility in its pathogenesis. Despite advances from deeper sequencing in larger groups, genome-wide approaches (GWAS) have traditionally focused on a “top-down” approach, with broad population-based discovery against clinical phenotypes (e.g., diabetes and obesity), generating a panoply of loci requiring functional characterization. While tissue-specific quantitative trait loci, epigenetics, and single cell tissue sequencing have begun to play a role in facilitating the functional interpretation of GWAS findings, aggregate phenotypes (e.g., body mass index, diabetes) do not capture the rich metabolic heterogeneity within obesity and dysmetabolism. Furthermore, identifying functionally relevant genes at gene-dense loci from GWAS remains elusive, challenging deep exploration of underlying mechanistic-functional consequences.

Given increasing interest around “action at a distance” by specific adipose tissue-derived mediators (which may be targetable) in obesity complications, exploring adipose-derived mediators in human studies may provide a functional framework for elucidating the molecular basis for GWAS findings, leading to new methodological and translational developments in genome-scale analysis of medically relevant phenotypes. Indeed, extracellular vesicles (EV)—50-100 nm lipid-bound circulating vesicles that carry molecular cargo across tissues—have been recently featured in dysmetabolic effects in obesity (e.g., insulin resistance, hepatic dysfunction, cancer^10–12^). Adipose tissue may be a significant source of circulating EVs in obesity^11^, prompting studies of EV content in obesity and their potential function^13–16^. Despite this rich emerging field, leveraging the power of modern multi-omics to decipher causal-functional implications of transcriptional cargo within EVs—specifically EV cargo dysregulated in the obese state and extruded from adipose tissue—remains completely unexplored.

Given our recent studies suggesting cellular transcriptional states may be reflected in RNA content of EVs released by source cells^17,18^, we hypothesized that transcriptional cargo from circulating EVs that characterize obesity would mirror adipose tissue transcriptional states that define causal-functional predisposition to obesity and related diseases. We first characterized circulating EVs in obese and lean humans, including transcriptional cargo differentially expressed in obesity. Given circulating EVs may have a wide origin across the human organism, we next compared this “obesity EV transcriptome” to expression from human visceral adipose tissue (VAT)-derived EVs from freshly collected and cultured biopsies from the same obese individuals. Using genes prioritized within EVs, we finally leveraged functional genomic approaches to interrogate adipose-specific regulation of gene expression reflected in the obesity EV transcriptome, (1) identifying genetic effects on obesity and across obesity-related metabolic-inflammatory diseases in a large-scale biobank and GWAS meta-analysis and (2) determining the phenotypic consequences of obesity-associated gene knockouts in published models. To our knowledge, the current work is the first to link tissue-relevant transcriptional states in human obesity sampled by EV transcriptomics to the richness of human genetic approaches for downstream prioritization of regulatory elements with target genes that may ultimately be targetable to prevent progression of metabolic disease.

## RESULTS

### Clinical populations and EV characterization

The overall study schema is shown in **Figure 1**. Baseline demographic and clinical characteristics of our analytical cohorts are shown in **Table 1**. By definition, obese cohorts had increased body mass index (BMI), predominantly in the moderate to severe obesity range (BMI > 35 kg/m^2^). The cohorts spanned a broad age and sex distribution, with a significant diabetes prevalence across samples. *Ex vivo* cultured visceral adipose tissue explants (VAT; from obese cohort) exhibited high protein expression of adipose tissue specific markers (adiponectin, perlipin1 and GLUT4) as well as morphologic and immunohistochemical characteristics of adipose tissue (by adiponectin immune reactivity; (**Supplementary Figure 1**). EVs isolated from lean individuals and circulating and *ex vivo* VAT-derived EVs (from culture) in obese individuals (obese cohort) demonstrated a particle number and size distribution (65-100 nm) which aligns with published morphometric parameters, though we observed slightly more EVs in plasma from obese individuals (**Figure 2A**; average obese plasma: 1.43×10^10^ conc/mL; lean plasma 5.08×10^9^ conc/mL; *ex vivo* VAT-derived EVs: 4.48×10^9^ conc/mL). Canonical EV surface markers (CD63, CD81) and cargo proteins (Alix, TSG101) were present across groups, while perilipin1—an adipose tissue biomarker^19^—was higher in plasma EVs from obese individuals and *ex vivo* VAT-derived EVs (also from obese individuals). We did not observe contamination by intracellular proteins (via 58K Golgi protein; **Figure 2B**). EVs exhibited a distinctive cup-shaped morphology encapsulated by a double-layered lipid bilayer membrane by transmission electron microscopy (**Figure 2C**).

**Figure 1.**
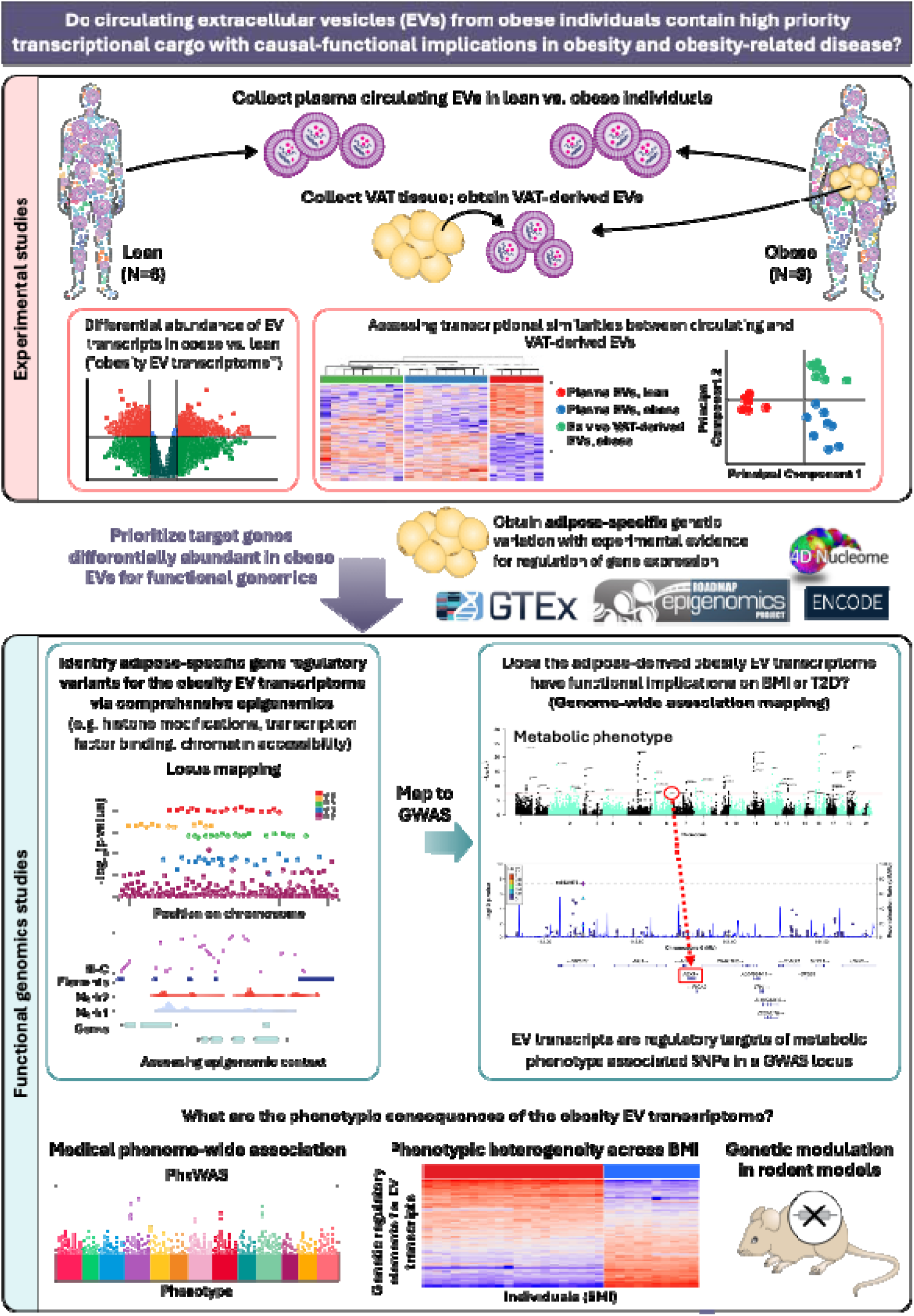
Study approach.

**Figure 2.**
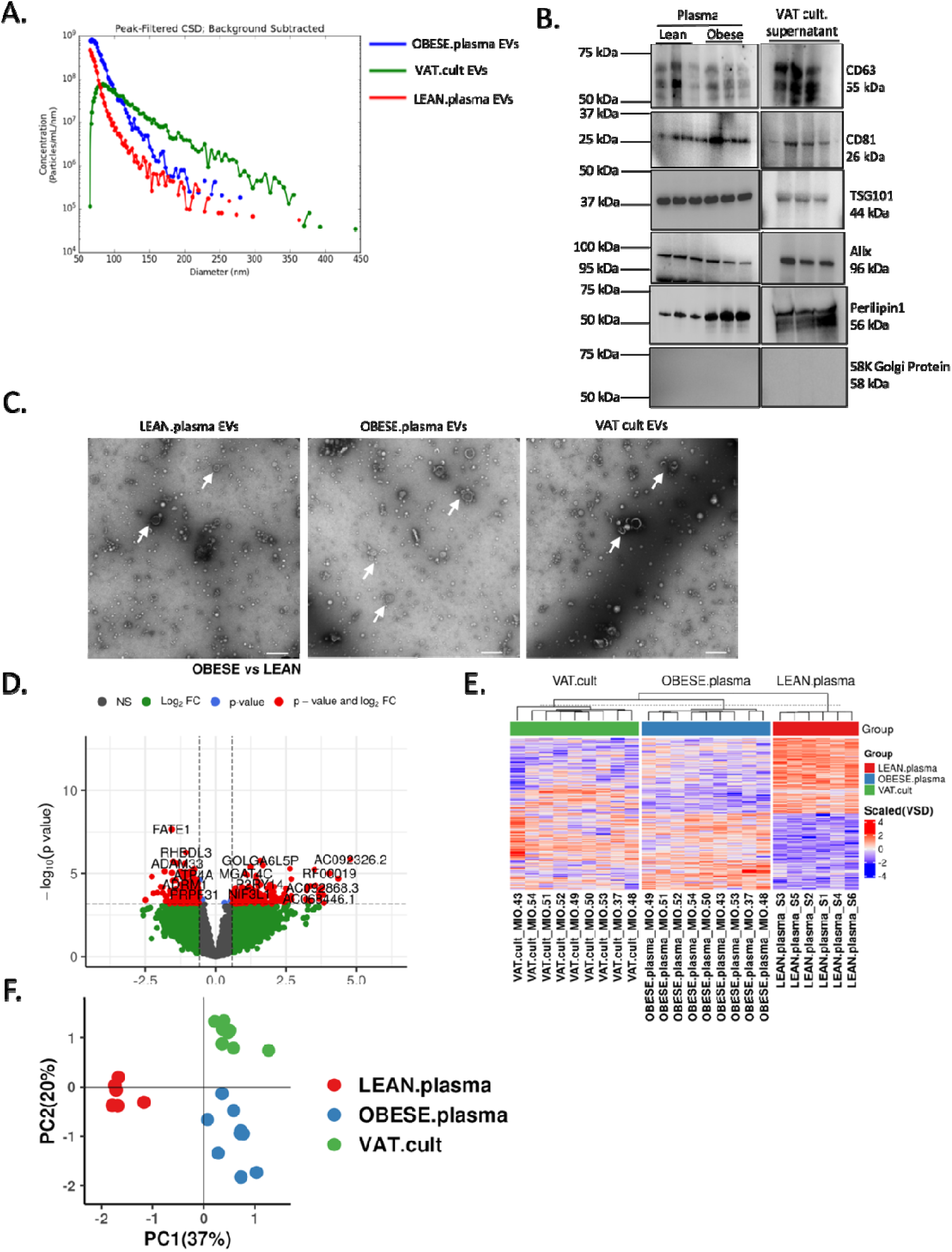
Adipose EVs and Plasma EVs share transcriptome during obesity. **A.** Representative microfluidic resistive pulse sensing showing concentration and size distribution profiles of the EV population isolated by SEC. **B.** Representative Western blot of the expression of CD63, CD81, TSG101, Alix, Perilipin 1, and 58K Golgi protein, as determined in the pooled EV samples from Lean, Obese plasma and VAT groups isolated by SEC. **C.** Isolated EVs were visualized using TEM (scale bar used = 500 nm). **D.** Volcano plots showing the significantly differentially expressed genes from the EV bulk RNA sequencing analysis in lean versus obese groups. Y axis shows Log_10_ p value and X axis displays the log_2_-fold change value. The red dots represent the differentially expressed genes with adjusted p value ≤ 0.1 and absolute_□_fold change ≥ 1.5, while green dots represent non significantly modulated genes. Positive log_2_-fold change means the gene is highly expressed in obese samples. **E.** Hierarchical clustering and **F.** PCA analysis of differentially expressed genes by group.

**Table 1.** Baseline characteristics of patient samples.

**Subject information for obese cohort**
| **Subject_ID** | **MIO.3** | **MIO.4** | **MIO.4** | **MIO.4** | **MIO_** | **MIO_** | **MIO_** | **MIO_** | **MIO_** |
| --- | --- | --- | --- | --- | --- | --- | --- | --- | --- |
|  | **7** | **3** | **8** | **9** | **50** | **51** | **52** | **53** | **54** |
| **Age range** | 35-40 | 60-65 | 20-25 | 35-40 | 35-40 | 25-30 | 40-45 | 50-55 | 40-45 |
| **Sex** | M | F | F | F | F | F | F | F | F |
| **Race** | Black | White | Asian | White | Not<br>listed | White | White | White | White |
| **BMI** | 50.87 | 46.02 | 41.86 | 55.09 | 40.58 | 48.07 | 45.93 | 49.26 | 40.69 |
| **BW (kg)** | 142.9 | 129.3 | 110.7 | 143.3 | 114.0 | 139.3 | 145.2 | 128.6 | 114.3 |
| **Height (cm)** | 167.6 | 167.6 | 162.6 | 161.3 | 167.6 | 170.2 | 177.8 | 160.0 | 167.6 |
| **Prediabetic/diabetic** | Y | N | N | Y | Y | N | N | N | N |
| **Plasma cholesterol** | 168 | No<br>lab | 162 | 172 | 141 | 199 | 177 | 190 | 144 |
| **Plasma HDL** | 38 | 46 | 44 | 42 | 44 | 34 | 37 | 68 | 47 |
| **Plasma LDL** | 11 | 112 | 100 | 108 | 84 | 116 | 118 | 103 | 67 |
| **Plasma triglycerides** | 93 | 93 | 90 | 109 | 66 | 245 | 112 | 90 | 149 |

Subject information for lean cohort
| Subject_ID | S1 | S2 | S3 | S4 | S5 | S6 |
| --- | --- | --- | --- | --- | --- | --- |
| Age range | 55-60 | 30-35 | 50-55 | 50-55 | 35-40 | 40-45 |
| Sex | M | M | M | M | F | F |
| Race | White | White | White | White | White | White |
| BMI (kg) | 22.96 | 21 | 18.6 | 23.7 | 22.08 | 29.03 |
| BW | 72.6 | 62.6 | 65.8 | 74.8 | 64 | 78.5 |
| Height (cm) | 177.8 | 172.7 | 188 | 177.8 | 170.2 | 163.8 |
| Prediabetic/diabetic | N | N | N | N | N | N |
| Plasma cholesterol | 191 | No lab | No lab | 146 | No lab | 177 |
| Plasma HDL | 66 | No lab | No lab | 53 | No lab | 66 |
| Plasma LDL | 113 | No lab | 79 | No lab | 100 | 67 |
| Plasma triglycerides | 62 | No lab | No lab | 72 | No lab | 54 |

| Subject_ID | Hs009 | Hs010 | Hs012 | Hs013 | Hs253 | Hs254 | Hs256 | Hs266 |
| --- | --- | --- | --- | --- | --- | --- | --- | --- |
| Age range | 40-45 | 30-35 | 35-40 | 20-25 | 50-55 | 40-45 | 40-45 | 65-70 |
| Sex | F | F | M | M | F | F | F | M |
| Race | Black | White | White | White | White | White | Black | White |
| BMI | 45.7 | 43.1 | 48.7 | 43.2 | 30.04 | 23.96 | 34.53 | 22.15 |

### Defining an obesity EV transcriptome in humans and its relation to VAT-derived EVs

Given that adipose tissue EVs may comprise a significant fraction of circulating EVs in obesity^11^, we sought to test whether transcriptional contents of circulating EVs that define obesity would be similar to that of specific VAT-derived EVs via broad transcriptomics^20^ (≈84% mapped to human genome, ≈6% of uniquely mapped reads assigned to genes). In an analysis that included <u>all expressed transcripts (minimum 5 median reads in any one group) across all groups</u>, we observed distinct clustering of plasma circulating EV transcripts from VAT-derived EVs (**Supplementary Figures 2A-B**) consistent with origin of EV transcripts in plasma from cellular compartments such as platelets and hematopoietic cells. Focusing on the differences between lean and obese observed in the PCA (**Supplementary Figure 2B**), we identified 282 transcripts (corresponding to 277 unique gene symbols) differentially expressed within circulating EVs between lean and obese individuals (at an absolute fold-change ≥ 1.5, 10% false-discovery rate; **Figure 2D**), dubbed the “obesity EV transcriptome.” We observed a strikingly similar overall pattern between plasma circulating EVs and VAT-derived EVs in obese individuals when we restricted our analysis to those 282 differential expressed transcripts (**Figure 2E-F**). Mapping the circulating EV transcripts from obese individuals to extant <u>matched</u> single cell/nuclear transcriptomic data derived from VAT tissue from the same individuals suggested a wide distribution of expression of those 282 differential expressed transcripts across multiple cell types relevant to human obesity (e.g., adipocytes, inflammatory cells; **Supplementary Figure 3A**).Overall, these results suggested that transcripts in circulating EVs from obese individuals—specifically those different from EVs in lean individuals—had a similar expression pattern to that of VAT-derived EVs, consistent with our hypothesis that differences between lean and obese EV contents would mirror the adipose tissue state.

### Comprehensive functional genomics of the obesity EV transcriptome enhances annotations within GWAS of obesity and diabetes

We next investigated the potential functional implications of the 277 unique genes corresponding to the differentially expressed EV transcripts through comprehensive genomic approaches across BMI and type 2 diabetes (T2D). We first identified adipose-specific *cis*-regulatory elements in (chromatin) contact with each corresponding gene using epigenomic, chromatin accessibility, and chromatin interaction assays (Methods). We stratified all SNPs (regardless of genome-wide significance) from high-powered GWAS of BMI (N=419,163) and T2D (428,452 cases and 2,107,149 controls) based on whether they overlapped one of these elements, annotating their “functional status”. Of the 13,791,467 SNPs tested for BMI, 61,114 (0.44%) were linked to a differentially expressed EV transcript using this approach. For T2D, 144,987 of the original 11,028,900 GWAS SNPs (1.31%) were in an adipose regulatory element for an EV-implicated transcript.

We next analysed the GWAS of BMI, where 50,839 SNPs (many in high linkage disequilibrium [LD]) reached genome-wide significance (*p* < 5 × 10^−8^), representing 366 independent loci. After filtering these SNPs by functional status as outlined above, we identified a subset of 667 variants that were in a regulatory element in adipose for an obesity-associated EV transcript, representing 53 independent loci (**Figure 3A**). Similarly, in the GWAS of T2D^21^, 49,852 SNPs (again, many in strong LD) reached genome-wide significance, representing 543 independent loci. Functional status filtering reduced this set to 553 SNPs, representing 155 independent loci (**Figure 3B**). Converging our ‘functional genomic’ approach with the GWAS data set revealed that adipose-tissue specific, regulatory SNPs for the obesity EV transcriptome strikingly mapped onto many of the strongest GWAS BMI and T2D signals, specifying the target genes involved in these associations and implicating the EV transcriptome functionally in both phenotypes.

**Figure 3.**
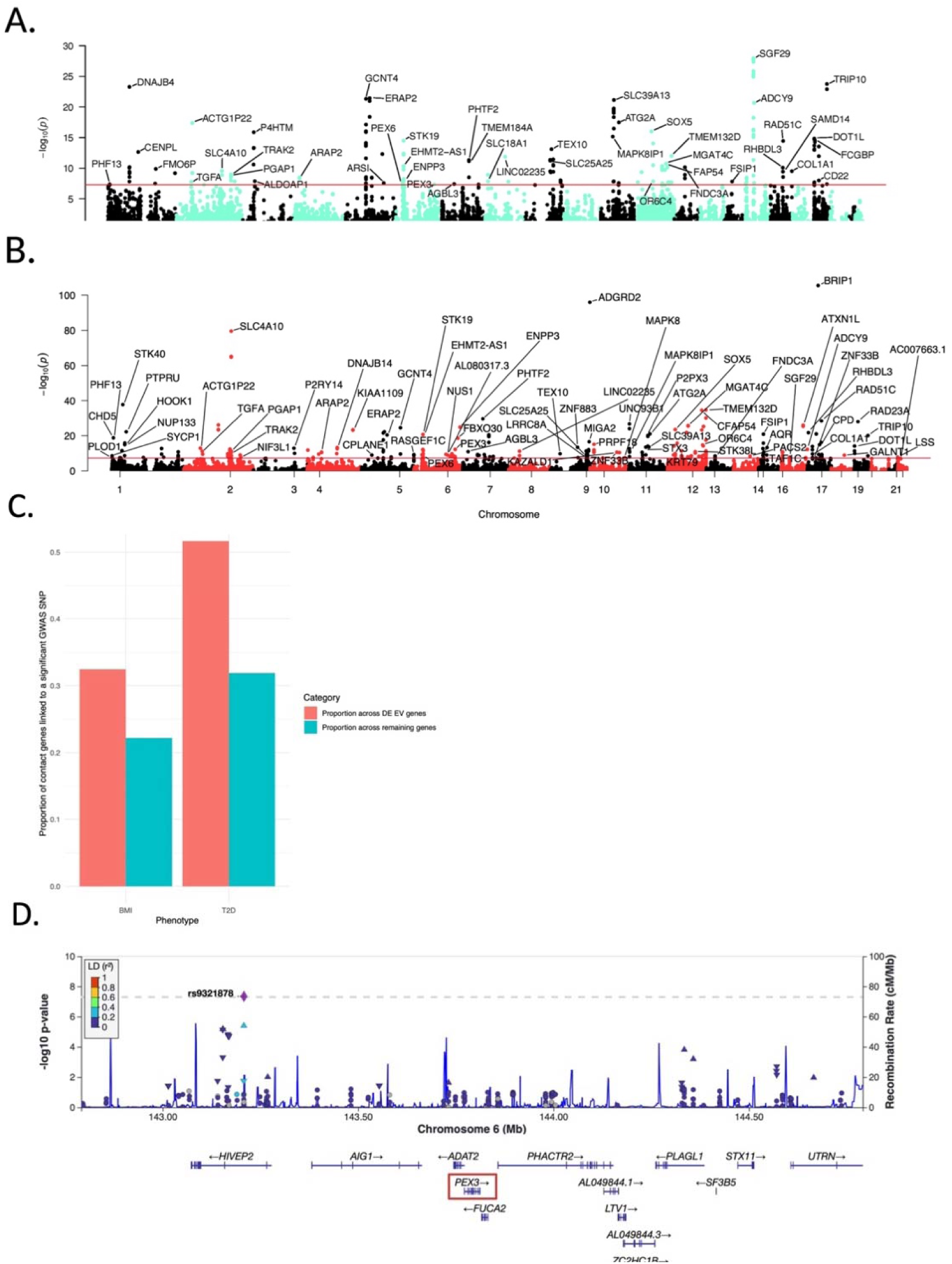
Adipose-specific regulatory elements of differentially expressed EV genes are enriched for GWAS SNPs associated with BMI and T2D. **A**. BMI GWAS (N = 419,163) associations for adipose-specific regulatory SNPs of the 282 differentially expressed EV transcripts. The red horizontal line denotes genome-wide significance (*p* < 5 × 10^−8^). The top SNP associated with each gene is labeled. **B.** T2D GWAS (N = 2,535,601) associations for adipose-specific regulatory SNPs of the 282 differentially expressed EV transcripts. **C.** Proportions of differentially expressed EV genes in contact (Hi-C) with an adipose regulatory element versus all remaining genes that can be linked to a BMI or T2D-associated SNP. **D.** SNP rs9321878, a variant within an intron of *HIVEP2* at locus 6q24.2. Hi-C contacts suggest that this SNP is in a regulatory element in contact with neighbouring gene *PEX3*, and knockout of *PEX3* in a murine model resulted in a classic metabolic phenotype.

Accordingly, the obesity EV transcriptome was highly enriched in GWAS-implicated, adipose-specific regulatory SNPs: for both BMI and T2D, we observed a significantly higher proportion of differentially expressed EV-implicated genes that were linked to a significant (*p* < 5 × 10^−8^) regulatory GWAS SNP relative to the rest of the genes with adipose-specific regulatory elements (**Figure 3C**). For BMI, 32% of differentially expressed EV genes with adipose regulatory-contact data were linked with a significant GWAS SNP, **Figure 3D**), versus 22% of the remaining genes (two-proportion z-test *p* = 2.48 × 10^−3^). For T2D, 52% of differentially expressed EV genes with adipose regulatory-contact data were linked with a significant GWAS SNP, versus 32% of the remaining genes (two-proportion z-test *p* = 1.99 × 10^−7^). These results provide evidence that transcripts within the obesity EV transcriptome were notably enriched for adipose-specific regulatory SNPs that were highly significant for associations with BMI and T2D. Our integrative analysis substantially enhanced regulatory annotations at GWAS-implicated loci for BMI and T2D.

### Phenome-wide association of regulatory SNPs for the obesity EV transcriptome is enriched for metabolic-inflammatory phenotypes physiologically linked to obesity

To further parse broad, phenotypic implications of obesity EV transcriptome regulatory elements from genomic studies, we performed a phenome-wide association study (PheWAS) in ≈500K individuals in the UK Biobank across 514 heritable traits^22^. This analysis utilized the SNPs within a regulatory element in adipose tissue for the obesity-associated adipose-derived EV transcripts described in the prior section. We identified 24,086 significant (*p* < 1.59 × 10^−9^) SNP-phenotype associations, representing 4,740 distinct SNPs. Among the top 50 traits associated with the regulatory SNPs for the EV-implicated genes, 17 were inflammatory phenotypes, with prominent enrichment for T2D and BMI. We also found a variety of autoimmune and inflammatory conditions with known comorbidity, worsened disease severity, or response with obesity, including celiac disease^23–24^, ulcerative colitis^25–26^, psoriasis^27^, asthma^28^, among many others (**Figure 4**). These results harness adipose-specific regulation of the obesity EV transcriptome to implicate EV cargo not only in direct complications of excess adiposity (obesity, diabetes), but also to a broader set of disease conditions with epidemiologic links to overweight/obesity.

**Figure 4.**
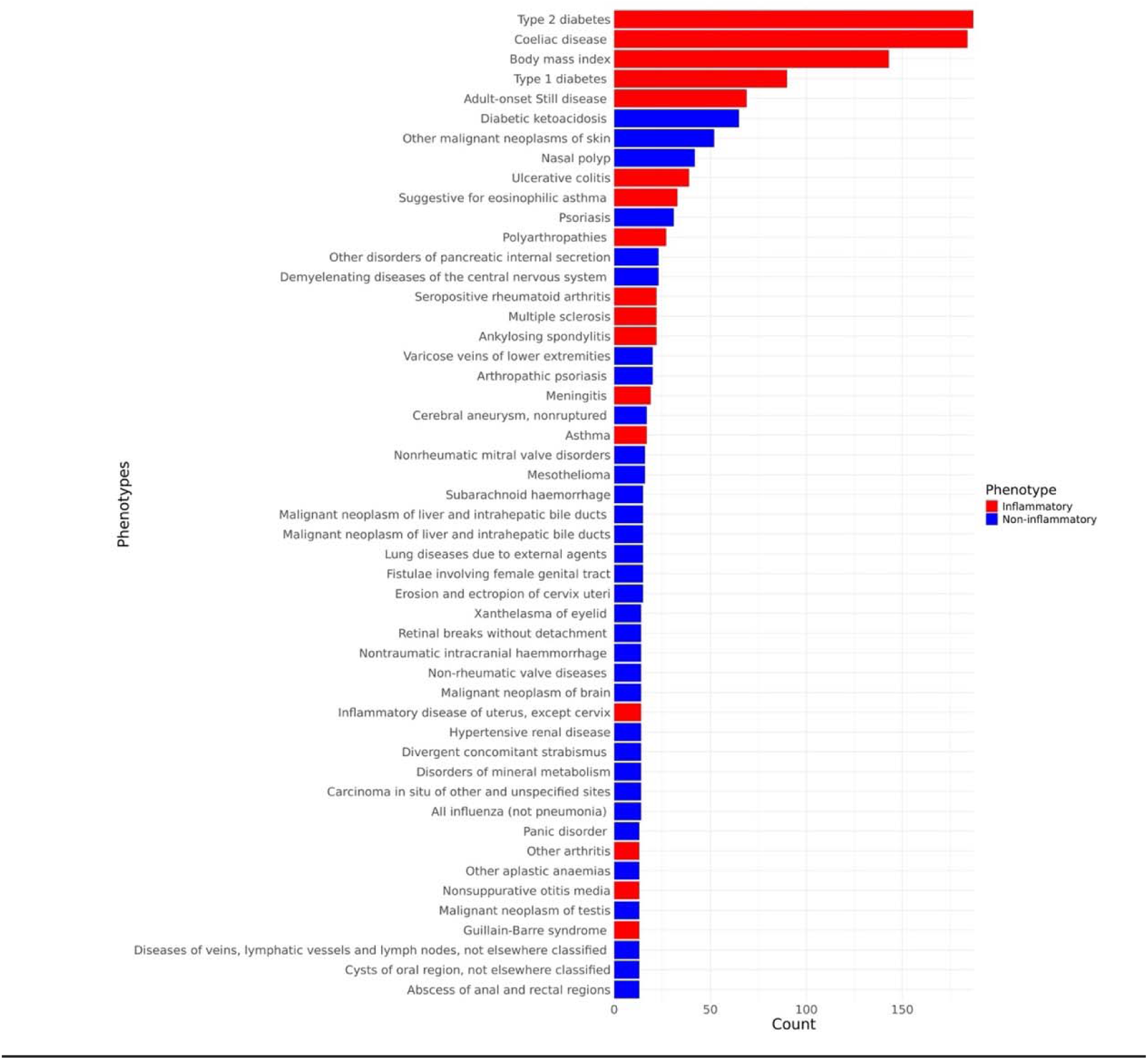
Phenotypic associations for SNPs overlapping an adipose-specific regulatory element in contact with one of the_282 differentially expressed EV transcripts. Among the top 50 traits associated with the regulatory SNPs for the EV-implicated genes, 17 were inflammatory phenotypes, with prominent enrichment for T2D and BMI. Traits labeled in red are inflammatory, while those in blue are non-inflammatory. Each phenotype shown here is implicated by a SNP association (*p* < 1.59 × 10^−9^) for a SNP in an adipose-specific regulatory element for an EV-implicated gene.

### The obesity EV transcriptome linked to BMI-associated regulatory SNPs demonstrate metabolic phenotypes in a model system

To provide another layer of validation for our functional genomics approach around adipose genes encoding obesity EV transcriptome, we next interrogated available murine models genetically altered for key implicated genes for metabolic phenotypes. Of the 282 transcripts within the obesity EV transcriptome, we identified 49 that were in contact with an adipose regulatory element that overlapped a significant BMI-associated SNP (*p* < 5 × 10^−8^). Of these 49 transcripts, 17 resulted in a phenotypic alteration when deleted in mice^1,2^, four of which resulted in a classic metabolic phenotype and plausible mechanisms of obesity-related metabolic dysregulation (*SGF29*, *FNDC3A*, *ADCY9*, *PEX3;* **Supplementary Table 2**). Some, but not all, functional consequences were directionally consistent with differential expression in EVs from obese versus lean individuals. For example, two of these genes were downregulated in circulating EVs from obese individuals (SAGA complex associated factor 29 [*SGF29*] and adenylate cyclase 9 [*ADCY9*]) and associated with significant alteration in glucose metabolism (decreased glucose level, increased insulin level, respectively); the other two were increased in circulating EVs from individuals with obesity (fibronectin type III domain containing 3A [*FNDC3A*] and peroxisomal biogenesis factor 3 [*PEX3*]) with mouse deletion resulting in effects on body composition and pro-atherogenic dyslipidemia or inflammation, respectively. Prior literature implicated several of these genes in obesity-relevant mechanisms but not directly in obesity, including *SGF29* (endoplasmic reticular stress response^29^), *PEX3* (redox balance and metabolic pathways^30^ and peroxisome biogenesis^31^), and *ADCY9* (context-dependent cardiovascular mechanisms^32^). Strikingly, our functional genomics approach to the obesity EV transcriptome resolved the gene of interest (1) within a gene-dense loci implicated by GWAS (e.g., *PEX3, SGF29*) or (2) in a locus where the SNP association is not proximally linked to the target (*FNDC3A*; **Supplementary Figures 4-6**). In addition, incorporation of the EV data clarified SNPs that may regulate target genes not in traditional *cis* linkage (e.g., ADCY9 in **Supplemental Figure 4B**). Effectively, the combined EV-functional genomics approach enhanced the functional interpretation of GWAS hits.

## DISCUSSION

While the systemic inflammatory clinical consequences of obesity are increasingly recognized, how metabolically active adipose tissue in obesity transduces its clinical expression remains actively debated. A key step in this understanding has been to define pathway-specific functional marks from adipose tissue and map these marks to human obesity and its related disorders. In this context, traditional genomic approaches have yielded a broad array of reproducible genomic locations (e.g., *FTO*^2^) associated with BMI, with downstream integration of “multi-omic” circulating (and increasingly tissue-based) human data to filter molecular signals most relevant to disease. While single cell adipose tissue sequencing has availed the community of complementary data, these methods remain invasive and difficult to deploy in a wider context (including in lean individuals). Extracellular vesicles have shown the potential to contain functionally active cargo that mediate metabolic effects across obesity-relevant organs and disease states (e.g., liver, muscle, diabetes, exercise), with enrichment in adipose tissue-derived EVs in obese states^11^. Moreover, the EV transcriptome (particularly mRNA and non-coding RNA fragments) appears to reflect key features of the transcriptome of the source cell. Nevertheless, leveraging EV data to guide a function-informed, intercellular communication driven genome-scale analysis has not been undertaken.

Here, using state-of-the-art EV characterization methods, we defined a differential EV transcriptome—the “obesity EV transcriptome”—across lean and obese states in humans, demonstrating the similarity in obesity-defining EV transcripts between bulk circulating vesicles and EVs derived from *ex vivo* culture visceral adipose tissue from the same individuals. Using a multimodal genomic approach that integrated adipose tissue-specific quantitative trait loci, epigenetic, and regulatory information, we found that regulatory elements for the transcripts in the obesity EV transcriptome strongly co-localized to top genomic loci associated with BMI and T2D in GWAS, providing specific gene annotation in gene-dense GWAS loci. Across ≈500K individuals in the UK Biobank, we demonstrated substantial enrichment of these obesity EV transcript-linked regulatory elements with metabolic-inflammatory conditions (including obesity and T2D as top hits). Genetic ablation of several targets in murine models recapitulated dysmetabolic phenotypes. Strikingly, EV transcripts that were different between obese and lean highlighted genes whose regulatory elements were colocalized with large effect size SNPs in a large-scale GWAS of BMI, thereby fine-mapping the functional genes at these GWAS-implicated loci. These results provide strong evidence underscoring the use of tissue-specific EV transcripts with broad genomic-epigenetic information to fine map functionally relevant targets in GWAS loci.

This work comes amidst an emerging precedent for EV cargo in mediating disease consequences. Increased EV secretion from adipose tissue and metabolic shift in EV cargo in response to obesity-induced ultrastructural tissue changes (e.g., inflammation, hypoxia) have been well demonstrated^33–35^, including an ability for EVs derived from obese models to transfer adverse metabolic changes to normal weight recipients^15–16, 36^. Elegant work has demonstrated the role of these adipose tissue-derived EVs to engage in cell-cell communication at organism-wide scale^37^ and across endocrine pathophenotypes (insulin resistance^10^) and pathways (insulin signaling, mTOR, among others^38^) central to obesity-related clinical risk. While the majority of circulating EVs arise from hematopoietic cells, immunodepleting circulating EVs of hematopoietic origin yields an EV fraction enriched for adipocyte markers^39^, suggesting active secretion of adipose tissue-derived EVs in circulation that may be expanded in obesity^40^. While several groups have postulated isolation of adipose tissue-specific EVs directly through canonical adipocyte proteins (e.g., FABP4^41^), these studies are in a nascent phase, with isolation of specific EV populations at scale a continued major limitation.

The critical innovation in this work involved two major steps. First, in addition to defining differences in transcript abundance between EVs from obese and lean volunteers, we isolated, cultured, and derived EVs from VAT *ex vivo* in the same individuals with obesity undergoing circulating EV characterization. While the lean and obese EV profile <u>across the entire transcriptome</u> were more similar to each other relative to the VAT-derived EVs, when <u>only transcripts differentially expressed within the lean versus obese states were considered</u>, circulating EVs from obese individuals were transcriptionally similar to the EVs derived from cultured VAT. This observation was consistent with prior work suggesting an increase in adipose tissue-derived EVs during obesity, leading to a conclusion that this “obese EV transcriptome” may serve as circulating marker of the transcriptional state of VAT in obese individuals. In recognition that these differences may be context-specific (dependent on populations studied and comorbidity), we next leveraged the power of human genetics to study the functional genomic implications of the obese EV transcriptome and its regulation in large populations.

Given our assertion that the obese EV transcriptome may be a hallmark of adipose tissue states, we predicated our human genetic approach on <u>adipose tissue-specific regulatory variants</u> for the obese EV transcriptome. This approach represents a fundamental departure from the usual pathway of genomic discovery in obesity and metabolic disorders: instead of predicating genetic discovery on anthropometric indices of obesity^2–3^, the current approach utilized circulating EV transcripts that may reflect both obesity and adipose tissue states to focus genomic studies of obesity and obesity-related disease liability. Importantly, we utilized an integrative multi-omics-based approach, based on 19 informative epigenomic, chromatin accessibility, and chromatin interaction assays^42^, to identify adipose-specific *cis-*regulatory elements for the transcripts in the EV obesity transcriptome. Our approach comprehensively exploits adipose-specific data on histone modifications (ChIP-seq), chromatin accessibility (ATAC-seq and DNase-seq), transcription factor binding (TF ChIP-seq), and chromosome conformation (Hi-C) to identify the regulatory elements to the EV-implicated genes, thus providing multiple levels of evidence for the regulatory hypothesis. These transcripts were then linked to GWAS-implicated SNPs based on overlap with these regulatory elements. PheWAS of these SNPs identified enrichment for inflammatory conditions, including both BMI and T2D. Notably, four of these EV transcripts linked to a BMI-associated regulatory SNP, when perturbed in a murine knockout model, resulted in a classic metabolic phenotype. These results collectively support the notion that adipose-derived EVs in obesity bear transcripts that have regulatory impact on broad, systems-wide metabolic phenotypes representing key obesity-related complications.

Several limitations to our study merit mention. We recognize that—while well characterized—our EV discovery cohort was small and at the extremes of obesity. Nevertheless, high concordance between circulating EVs and VAT-derived EVs (among genes within the obesity transcriptome) as well as differential expression and downstream genetic validation studies provide biological support. Certainly, evaluation across a broader patient population with more diverse characteristics (including external factors, metabolic function, and genetic background) is warranted. In addition, while genetic and epigenetic mapping identified adipose-specific functional elements in a normative context (not necessarily in tissues from obese individuals), the validation of these elements in PheWAS provides strong support of their plausibility. Functional validation of all identified targets via knock-out studies was outside the purview of the current report, though we envision that these studies provide a translational platform to harness tissue-specific EV RNA expression (as well as other molecular species) alongside functional genomics to identify novel targets to interrupt metabolic disease.

In summary, our findings feature EV transcripts dysregulated across a clinically important, targetable condition (obesity) as a substrate for causal-functional genetic discovery. By identifying an “obesity transcriptome” that reflects tissue states in humans and mapping functional genetic control of these transcripts using the power of modern genomics, we provide human translational evidence to support the idea that adipose-derived EVs in obesity harbor transcripts that may impact broad metabolic states in humans, including the complications of obesity. The presented approach represents a new paradigm in functional genomics at the nexus of genome and epigenome-wide human genetic annotations and EV transcription, enabling powerful future genomic studies of complex diseases.

## METHODS

### Study population and cohort procedures

The overall flow of the experimental approach is shown in **Figure 1**. Our study included samples from three cohorts. In our obese cohort (“Obese plasma EV and *ex vivo* VAT cohort”), we recruited 9 obese subjects undergoing bariatric surgery to obtain peripheral venous blood and VAT tissue during surgery as part of the EVOC study (<u>NCT06408961</u>). In lean cohort (“Lean plasma EV cohort”), we collected blood samples from 6 lean subjects. In our snRNA seq cohort (“Obese snRNA-seq and EV cohort”), we collected serum samples from 8 obese patients (**Supplementary Figure 3A**). Obesity was defined as a body mass index (BMI) greater than or equal to 30 kg/m^2^ whereas, BMI in lean samples was less than or equal to 25 kg/m^2^. Clinical and demographic data were systematically collected from all participants, as comprehensively outlined in **Table 1** to facilitate subsequent analyses and correlations with experimental outcomes.

Across all cohorts, we isolated plasma within 60 minutes of venipuncture via centrifugation as described previously^43^ and stored plasma at -80°C until EV isolation. Blood was collected from 8 subjects of the snRNA seq cohort in SST blood collection tubes and serum fraction was collected as described previously^44^. Aliquots of cell-free serum were stored immediately at −80 °C for future study.

### *Ex vivo* VAT explant culture

A small segment of visceral fat, concomitant with each “Obese plasma” sample was collected from the greater omentum of 9 obese patients from obese cohort during surgery and adipose explants were cultured adhering to protocols described in earlier studies with some modifications ^45–46^. Briefly, visceral adipose tissue was minced into small pieces with scalpel and plated into culture dishes with serum free conditioned media for 48 h. The explant derived media was centrifuged at 2000 × g for 10 min and supernatant was filtered using 0.8μM syringe filter to avoid any cellular debris contamination. Filtered visceral adipose tissue media (VAT) was stored at -80°C for later use.

### EV isolation

For isolation of EVs, the collected media was concentrated up to 510μl using 10kda column (Amicon Ultra-15 Centrifugal Filter Unit, EMD Millipore cat# UFC900308). Then EVs from plasma and cultured media were isolated using SEC-based Izon technology (Izon Science) as previously described^43^. Fractions 7–10 were pooled and used for downstream experiments as optimized by our group^43^.

### Microfluidic resistive pulse sensing (MRPS)

To measure size distribution and particle number MRPS analyses were performed. EVs were diluted at 1:100 to prevent saturation of the upper limit of detection or aggregation, subjected to MRPS using the Spectradyne’s nCS1 (Spectradyne, Signal Hill, CA, USA), and analysed with both high and low sensitivity settings (NP100; voltage, 0.60 V; stretch, 46.0 mm and NP400; voltage, 0.40 V; stretch, 43.5 mm respectively). The pressure was preset at 7.0 mbar. Minimum 2000 particles were analyzed for each sample.

### Transmission electron microscopy (TEM) of plasma and VAT EVs

Purified EVs were visualized using transmission electron microscopy (TEM). A 5 μL drop containing EVs was placed onto parafilm, and a glow-discharged, carbon-coated copper grid was placed on top for 30 minutes. Glow discharge treatment (30 seconds) rendered the grid surface hydrophilic. Grids were stained with 0.75% uranyl formate for 1 minute. EVs were visualized using a JEOL 1400 transmission electron microscope equipped with an Orius SC1000 CCD camera (Gatan, Inc. Pleasanton, CA, USA).

### Immunoblot

Western blot analysis was done as described, we have reproduced these methods with minimal modification from our prior work to allow scientific rigor and reproducibility^47^. Briefly, VAT and concentrated EV suspensions from plasma and culture media were lysed for protein extraction (RIPA lysis buffer; 1X protease and phosphatase inhibitor cocktail, Thermo Fisher Scientific) for 20 minutes at 4°C. Protein concentration was quantified with Pierce BCA Protein Assay Kit (Thermo Fisher Scientific) followed by SDS-PAGE. Gels were transferred to PVDF membranes (MilliporeSigma) and blocked with 5% bovine serum albumin (Millipore Sigma) for 1 hour at room temperature. Primary antibodies against CD81, CD63, Alix, TSG101, 58K Golgi protein, Adiponectin, Perlipin1 and GLUT4 were incubated at 4°C overnight at 1:1,000 concentration (**Supplementary Table 1**) followed by incubation with secondary HRP-antibodies for 1 hour at room temperature. Blots were developed using the Super Signal Femto developer (Thermo Fisher Scientific).

### Histology (H&E staining and Immunohistochemistry)

Adipose tissue sections were fixed with 4% PFA and stained with either H&E or with the primary antibody like Adiponectin followed by incubation with Alexa Fluor 488-labeled secondary antibodies (Molecular Probes) to visualize under microscope. Antibodies incubated slides were mounted with Vectashield (with DAPI [4′,6-diamidino-2-phenylindole]) (Vector Laboratories, Newark, CA), the slides were examined under a fluorescence microscope (BioRad).

### Extracellular RNA isolation and sequencing

Extracellular RNAs (ExRNAs) were isolated from plasma and *ex vivo* VAT culture media of total 24 subjects in our obese and lean cohorts (Lean plasma, n=6; Obese plasma, n=9, *ex vivo* VAT culture, n=9) using the exRNeasy Serum/Plasma kit (QIAGEN, Germantown, MD, USA) as per manufacturer’s protocol, cDNA libraries were constructed using the SMARTer Stranded Total RNA-Seq Kit v2 Pico Input Mammalian (Takara Bio, San Jose, CA, USA) and sequenced using NextSeq 2000 platform (Illumina, San Diego, CA, USA).

### RNA sequencing analysis

Reads were trimmed to remove adapter sequences using Cutadapt (v4.8). Quality control on both raw reads and adaptor-trimmed reads was performed using FastQC (v0.12.1)(www.bioinformatics.babraham.ac.uk/projects/fastqc). Reads were aligned to the Gencode GRCh38.p13 genome using STAR (v2.7.11a)^48^. Gencode v38 gene annotations were provided to STAR to improve the accuracy of mapping. featureCounts (v2.0.6)^49^ was used to count the number of mapped reads to each gene. Genes on mitochondrial chromosome (chrM) were removed from the down-stream analysis. ComplexHeatmap was used for cluster analysis and visualization. Significantly differential expressed genes with absolute fold change >= 1.5 and FDR-adjusted p value <= 0.1 were detected by DESeq2 (v1.42.1)^50^. For each differential expression comparison, low expressed genes with less than 5 median read count in both conditions were excluded.

Adipose tissue snRNA data was downloaded from SCP1376 (parent reference^44^). We excluded one sample from the provided data as it had subcutaneous but not visceral fat (VAT). Our final included dataset contained 8 SAT samples and 8 corresponding VAT samples. The cells were classified as SAT/VAT across specific cell types and used to visualize the expression of the differentially expressed EV genes.

### Identification of adipose regulatory elements in contact with the gene for an obesity-associated EV transcript

We developed a regulatory annotation framework using a broad set of 18 epigenetic assays, including histone ChIP-Seq (for chromatin marks H3K27ac, H3K27me3, H3K4me1, H3K4me2, H3K4me3, H3K36me3, H3K79me2, H3K9ac, H3K9me3, and H4K20me1), chromatin accessibility (ATAC-Seq and DNase-Seq), and transcription factor ChIP-seq (for RNA polymerase II subunit POL2RA and cohesion complex subunits RAD21 and SMC3), in addition to transcription start^42^. Application of these epigenomic and chromatin accessibility data to adipose-derived biosamples from the EpiMap^51^ repository identified genome-wide adipose-specific regulatory elements. We further utilized 4D Nucleome project^52^ chromatin conformation capture (3C) Hi-C data at a 1 kb resolution from the RUES2 cell line (accession 4DNFITDFDM7G) to identify chromatin interactions in element-promoter regions for additional support for the epigenetic-assay-derived regulatory relationship between an element and a target gene. Briefly, the Hi-C contact matrix, in mcool format, had undergone processing with the gold standard 4D Nucleome pipeline (https://data.4dnucleome.org/resources/data-analysis/hi_c-processing-pipeline) and normalization using the iterative correction and eigenvalue decomposition (ICE) algorithm^53^. Cooler (v0.8.2)^54^ was used to export contacts, and the resulting contact matrix was converted to bedpe format. Contacts were filtered to retain contact pairs in which the respective contact regions overlapped with a regulatory element and one of the obesity-associated EV genes. These data allowed us to identify adipose-specific regulatory elements in contact with the 282 genes for the EV transcripts found to be differentially expressed between obese and lean individuals.

### GWAS SNP functional characterization using adipose-specific regulatory elements

We leveraged summary statistics from a GWAS of BMI (accession 21001) performed in individuals of European ancestry in the U.K. Biobank^22^ and a multi-ancestry GWAS meta-analysis of T2D^21^. The GWAS of BMI included 419,163 individuals, while the GWAS of T2D included 2,535,601 individuals (428,452 T2D cases and 2,107,149 controls). From the complete set of genome-wide significant (*p* < 5 × 10^− 8^) SNPs, those within an adipose-specific regulatory element in chromatin contact with a gene for an obesity-associated EV transcript were identified. Manhattan plots were generated using the qqman (v0.1.8) R package^55^. To statistically test differences in proportion of differentially expressed EV-implicated genes that were linked to a significant (*p* < 5 × 10^−8^) regulatory GWAS SNP in comparison with the rest of the genes with adipose-specific regulatory elements, a two-proportion z-test was utilized.

### Visualizing the genomic and epigenomic landscape of regulatory SNPs for obesity-associated EV transcripts

A LocusZoom plot showing regional visualization of association results relative to genomic coordinates and the positions of genes in the region was plotted using the LocusZoom.js interactive visualization tool^56^.

Genomic coordinates in the bigWig files from the EpiMap Repository were lifted over from genome build hg19 to hg38 using CrossMap (v0.6.5)^57^. The adipose-specific regulatory elements (from the regulatory annotation framework), element-gene contacts (based on Hi-C), BMI-associated SNPs (from the UK Biobank GWAS) in contact with an the gene for an obesity-associated adipose-derived EV transcript, and relevant chromatin marks were visualized relative to nearby genes using the Integrated Genome Viewer (IGV v2.18.4)^58^.

### PheWAS of SNPs in an obesity-associated gene regulatory element in adipose tissue

For all SNPs within an adipose-specific regulatory element in contact with the gene for an obesity-associated EV transcript, we performed a phenome-wide association study (PheWAS) across 513 additional heritable phenotypes in the U.K. Biobank^22^. SNP-phenotype associations with *p* < 1.59 × 10^−9^ (0.05 / 31,351,482 tested SNP-phenotype associations) were considered significant. Counts of significant SNP-phenotype association were plotted using the ggplot2 (v3.5.0) R package.

### Identifying phenotypic consequences of obesity-associated gene knockouts in mice

From an initial set of EV transcripts differentially expressed between obese and lean individuals and linked to a regulatory element harboring significant (*p* < 5 × 10^−8^) SNP associations with BMI, we identified a subset that showed a significant phenotypic change in their corresponding mouse knockout from the International Mouse Phenotyping Consortium (www.mousephenotype.org)^59^.

Study approval. The study was IRB-approved at Mass General Brigham, with written informed consent collected from participants before study initiation. The trial is registered at clinicaltrials.gov (<u>NCT06408961</u>).

## Data Availability

RNA-Seq data are available at the NCBI dbGaP (in process).

## Author contributions

EC conducted EV experiments. MJB, QS, and PL did statistical and computational analysis. GL conducted selected experiments. MPE, WBL, ORW, KL, CA, DG, MH, and KF collected and provided clinical samples and clinical metadata. KA, PS, MGC, PG, CRF, DY, JB, DY, ER, and KVJ provided critical review of the manuscript. EC, MJB, QS, ERG, RS, and SD participated in writing the manuscript. ERG, RS and SD were responsible for supervision of data analysis and for the final manuscript.

## Acknowledgements

This work was funded by grants from American Heart Association Strategically Focused Research Network (AHA; SFRN16SFRN31280008); National Heart, Lung, and Blood Institute (1R35HL150807-01); and National Center for Advancing Translational Sciences (UH3 TR002878) to SD. RS is supported by the NIH and the AHA. GL is supported by AHA (23CDA1045944). ERG is supported by the National Human Genome Research Institute (R35HG010718 and R01HG011138), National Institute of General Medical Sciences (R01GM140287), and the National Institute of Diabetes and Digestive and Kidney Diseases (U01DK140952). EDR is supported by the National Institute of Diabetes and Digestive and Kidney Diseases (RC2 DK116691). RS has served for a consultant for Amgen, Cytokinetics, and Thryv Therapeutics (with equity in Thryv). RS is a co-inventor on a patent for ex-RNAs signatures of cardiac remodeling and a pending patent on proteomic signatures of disease.

