## Supplementary Figures for "The extracellular vesicle transcriptome provides tissue-specific functional genomic annotation relevant to disease susceptibility in obesity"

**Supplementary Figure 1. Characterization of visceral adipose tissue explants. A**. Representative Western blot of the expression of Adiponectin, GLUT4 and Perilipin 1 in adipose tissue. **B.** Representative brightfield (H/E) and fluorescent images of the adipocytes (scale bar = 100 μm). DAPI (nuclear) and green (adiponectin) stains are shown. **C.** Schematic representation of adipose explants culture**.**

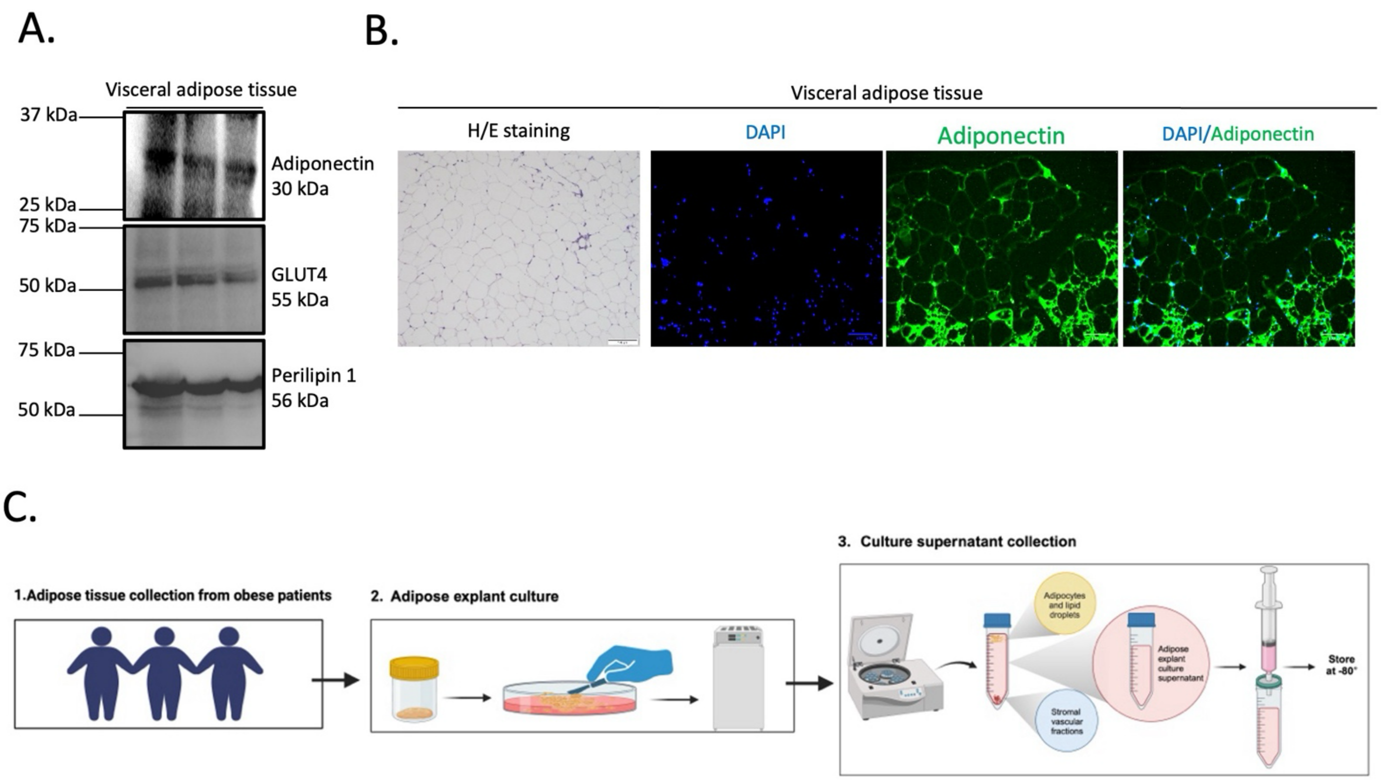

**Supplementary Figure 2. Bulk RNA sequencing of EV transcriptomes from obese and lean cohorts. A.** Hierarchical clustering of EV bulk RNAs by group in the obese and lean cohorts. B. PCA analysis of all genes from lean, obese plasma and VAT EV groups in obese and lean cohorts.

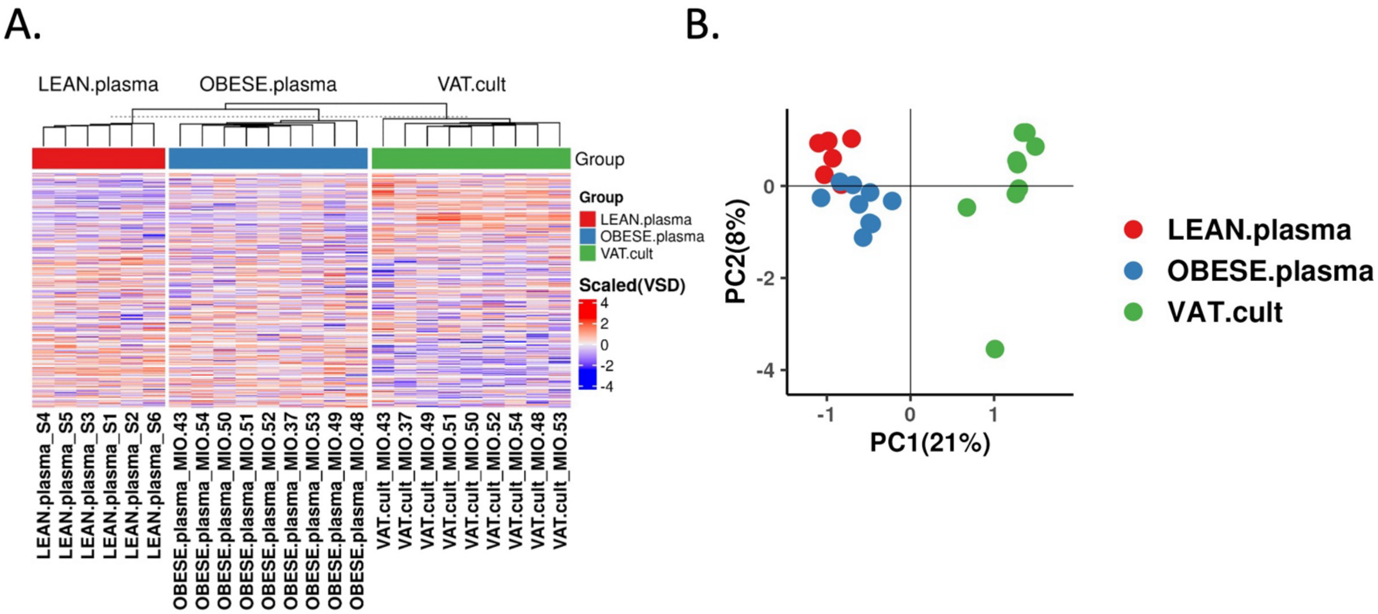

**Supplementary Figure 3. Adipose tissue transcripts are prevalent in circulation. A.** Dot plot analysis showing differentially expressed genes (obese versus lean plasma) that are present in the serum of obese patients are expressed across different cell types of adipose tissue with diverse expression (single-nuclear RNA sequencing) derived from the matched adipose tissue samples. Source of snRNA-seq described in text.

A.

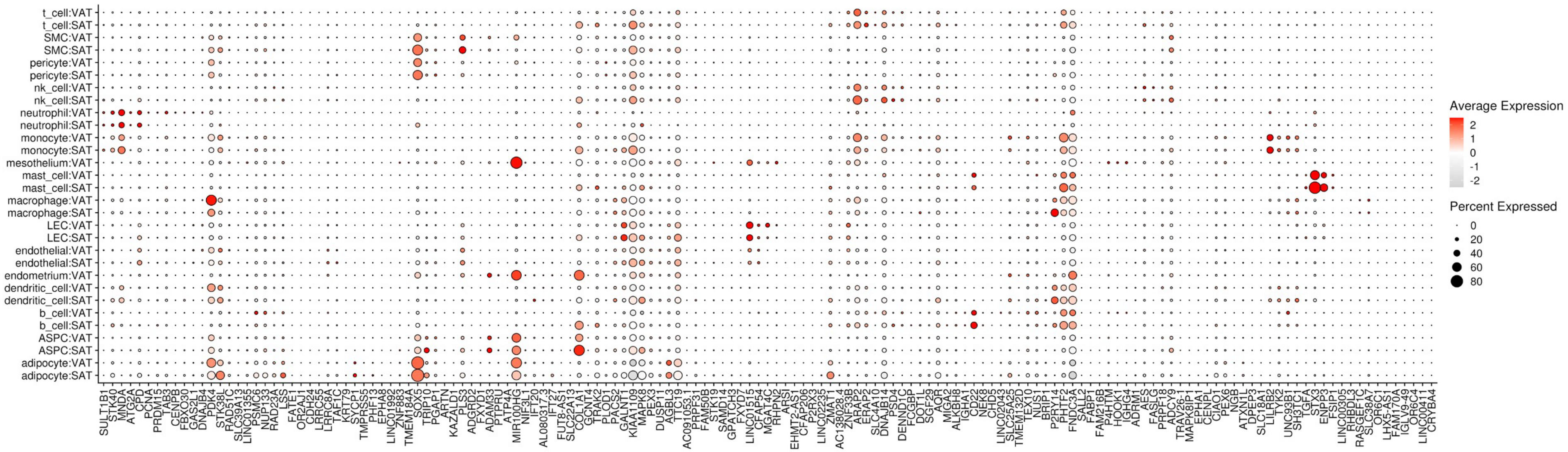

**Supplementary Figure 4. LocusZoom plots of BMI-associated SNPs linked to differentially expressed EV genes with mouse validation. A.** SNP rs3785354, a regulatory SNP of *SGF29*, at locus 16p11.2. This SNP is upstream of its target gene. SNP LD patterns are based on a set of European ancestry reference individuals. **B.** SNP rs17055384, a regulatory SNP of *FNDC3A*, at locus 13q21.1. This SNP is in an intergenic region 9 Mb downstream of *FNDC3A,* suggesting that it is a distal regulatory variant. **C.** SNP rs11643872, a regulatory SNP of *ADCY9*, at locus 16p11.2. This SNP is in an intron of *SETD1A* and is located over 26 Mb downstream of *ADCY9,* suggesting that it is a distal regulatory variant. **D.** SNP rs754611720, a regulatory SNP of *ADCY9*, at locus 16p13.3. This SNP is located directly downstream of *ADCY9.* **E.** SNP rs9321878, a regulatory SNP of *PEX3*, at locus 6q24.2. This SNP is within an intron of *HIVEP2,* a neighbouring gene of *PEX3*.

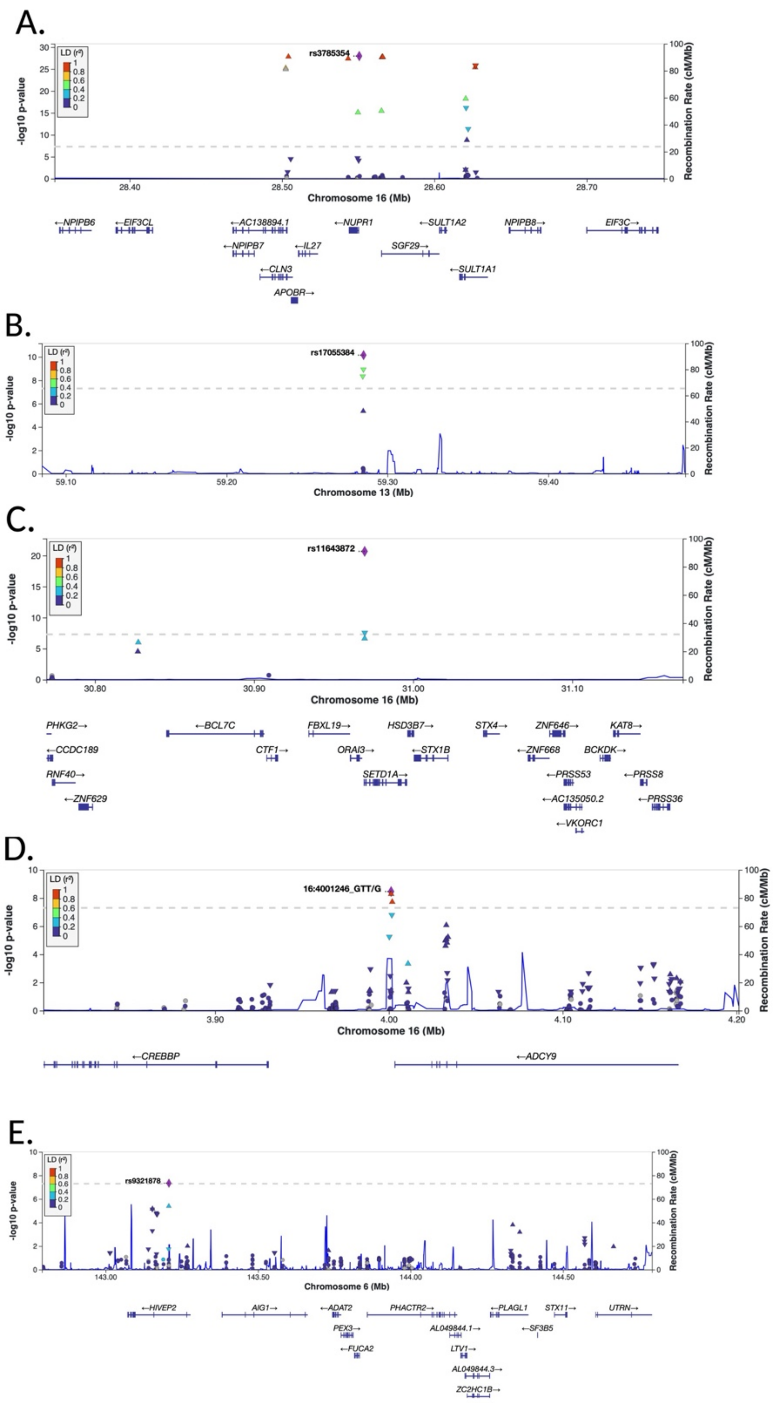

A.
rs3785354
LD (r²)
-log10 p-value
Recombination Rate (cM/Mb)
Chromosome 16 (Mb)
←NPIPB6
←EIF3CL
←AC138894.1
←NUPR1
←SULT1A2
NPIPB8→
EIF3C→
←NPIPB7
←IL27
SGF29→
←CLN3
←SULT1A1
APOBR→
B.
rs17055384
Chromosome 13 (Mb)
C.
rs11643872
Chromosome 16 (Mb)
PHKG2→
←BCL7C
FBXL19→
HSD3B7→
STX4→
ZNF646→
KAT8→
←CCDC189
CTF1→
ORAI3→
←STX1B
←ZNF668
BCKDK→
RNF40→
SETD1A→
←PRSS53
←PRSS8
←ZNF629
←AC135050.2
←PRSS36
←VKORC1
D.
16:4001246_GTT/G
Chromosome 16 (Mb)
←CREBBP
←ADCY9
E.
rs9321878
Chromosome 6 (Mb)
←HIVEP2
AIG1→
←ADAT2
PHACTR2→
←PLAGL1
STX11→
UTRN→
PEX3→
AL049844.1→
←SF3B5
←FUCA2
LTV1→
AL049844.3→
ZC2HC1B→

**Supplementary Figure 5. Chromatin contacts around BMI-associated SNPs linked to differentially expressed EV genes with mouse validation. A.** SNP rs3785354 is variant that is immediately upstream of *NUPR1* at locus 16p11.2. It overlaps with an adipose regulatory element that is in contact with *SGF29*. Contacts are derived from Hi-C in RUES2. **B.** SNP rs17055384 (grouped with the depicted SNP rs9569942) is an intergenic SNP at locus 13q21.1 that is approximately 9 Mb downstream of *FNDC3A.* However, Hi-C contacts indicate that this SNP overlaps an adipose regulatory that is in contact with *FNDC3A*. **C.** SNP rs11643872, a regulatory SNP of *ADCY9* at locus 16p11.2. Despite being over 28 Mb downstream of *ADCY9*, Hi-C contacts suggest that rs11643872 overlaps an adipose regulatory element in contact with this gene. **D.** SNP rs754611720, a regulatory SNP downstream of *ADCY9*, at locus 16p13.3. **E.** SNP rs9321878, a variant within an intron of *HIVEP2* at locus 6q24.2. Hi-C contacts suggest that this SNP is in a regulatory element in contact with neighbouring gene *PEX3*.

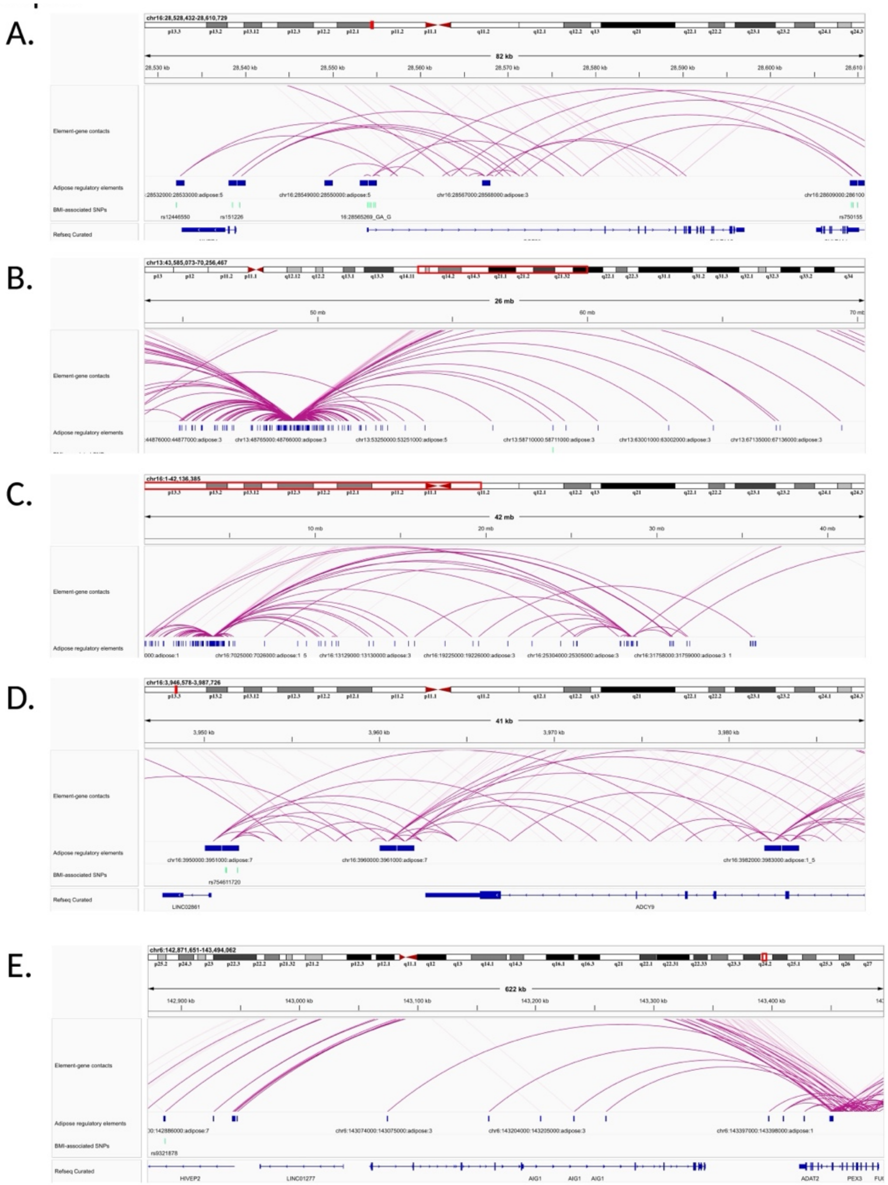
A.
B.
C.
D.
E.
82 kb
26 mb
42 mb
41 kb
622 kb
Element-gene contacts
Adipose regulatory elements
BMI-associated SNPs
Refseq Curated
ADCY9
HIVEP2
LINC01277
AIG1
ADAT2
PEX3

**Supplementary Figure 6. Epigenomic context around BMI-associated SNPs linked to differentially expressed EV genes with mouse validation. A.** SNP rs3785354, a regulatory SNP of *SGF29*, at locus 16p11.2. This SNP overlaps an enhancer-like promoter in adipocytes, characterized by EP300, H3K27ac, H3K4me3, and H3K9ac. **B.** SNP rs17055384, a regulatory SNP of *FNDC3A*, at locus 13q21.1. This SNP overlaps a transcribed element in subcutaneous adipose tissue, characterized by H3K36me3, H3K79me3, and H4K20me1. **C.** SNP rs11643872, a regulatory SNP of *ADCY9*, at locus 16p11.2. This SNP overlaps an enhancer-like promoter in adipocytes. **D.** SNP rs754611720, a regulatory SNP of *ADCY9*, at locus 16p13.3. This SNP overlaps an active enhancer in adipocytes, characterized by H3K27ac and H3K4me1. **E.** SNP rs9321878, a regulatory SNP of *PEX3*, at locus 6q24.2. This SNP overlaps a proximal insulator in adipocytes, characterized by CTCF, RAD21, and SMC3.

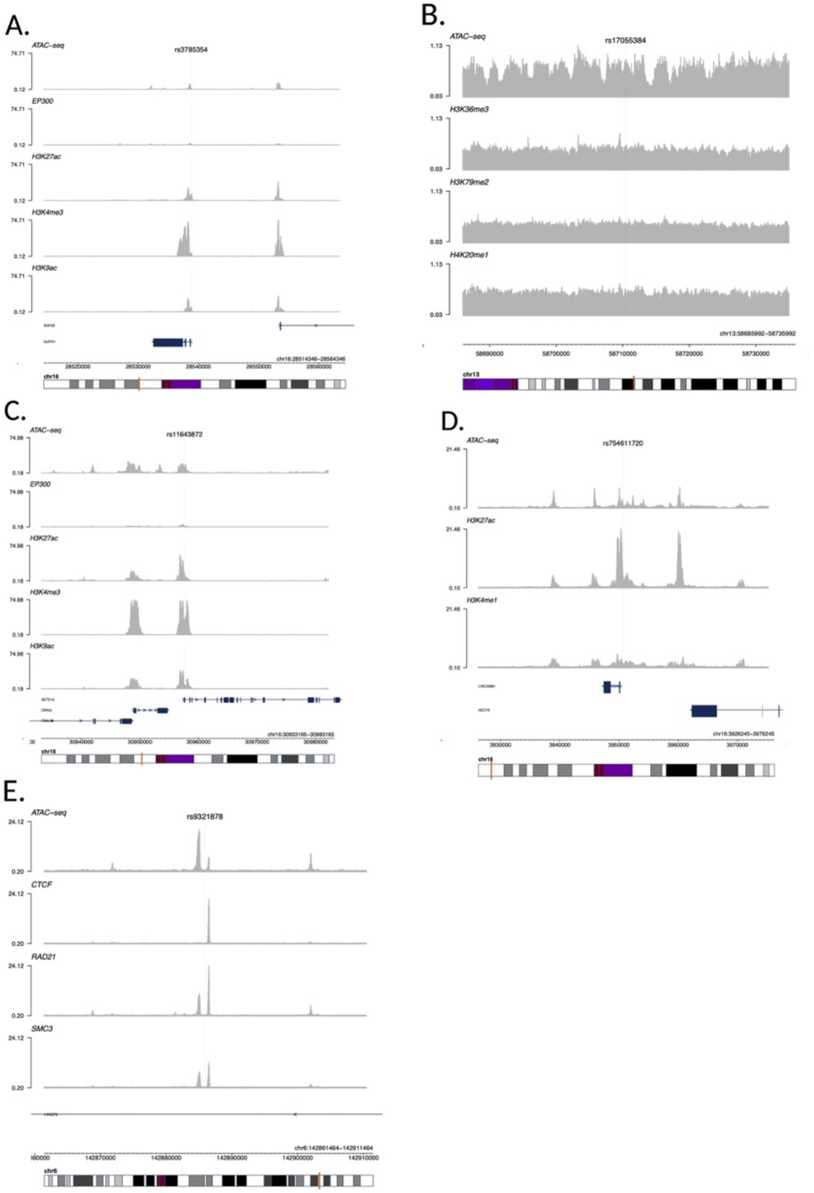

A.
ATAC-seq
rs3785354
EP300
H3K27ac
H3K4me3
H3K9ac
B.
ATAC-seq
rs17055384
H3K36me3
H3K79me2
H4K20me1
chr13
C.
ATAC-seq
rs11643872
EP300
H3K27ac
H3K4me3
H3K9ac
chr16
D.
ATAC-seq
rs754611720
H3K27ac
H3K4me1
chr16
E.
ATAC-seq
rs9321878
CTCF
RAD21
SMC3
chr6
